# Association Between Intermittent Water Supply and Helicobacter pylori Prevalence: A Global Ecological Study

**DOI:** 10.64898/2026.06.22.26356253

**Authors:** Saeed S Graham, Christin Wilkinson, Briannae Theodore, Mary-Jane Williams, Dmitry Tumin

## Abstract

**Background:** Helicobacter pylori is a major global pathogen with recognized potential for waterborne transmission. Intermittent water supply affects over one billion worldwide and may promote H. pylori contamination of municipal sources. Whether water supply discontinuity contributes to population-level H. pylori burden has not been examined globally.

**Materials and Methods:** We conducted a cross-sectional ecological analysis of 79 countries with matched utility-level water infrastructure data and country-level H. pylori prevalence estimates from a published global meta-analysis. The primary exposure was continuity of water supply (hours/day). Secondary exposures included non-revenue water percentage (NRW %), pipe breaks per utility, and operating cost coverage ratio. Unadjusted and adjusted linear regression models with heteroscedasticity-consistent standard errors were estimated, controlling for basic sanitation coverage and log-transformed population density. A sensitivity analysis used a population-based measure of water availability on demand.

**Results:** Greater water supply continuity was independently associated with lower H. pylori prevalence in both unadjusted (β = −0.987, 95% CI −1.669 to −0.305, p = 0.005) and adjusted models (β = −1.125, 95% CI −1.876 to −0.375, p = 0.004). Higher NRW % and lower operating cost coverage were each associated with higher H. pylori prevalence after adjustment. Pipe breaks were not significant in regression models though the Spearman correlation was in the expected direction. Sensitivity analysis produced consistent findings.

**Conclusion:** IWS and broader water infrastructure deterioration are associated with higher H. pylori prevalence at the country level. These findings implicate water supply continuity as a potentially relevant environmental determinant of H. pylori transmission and suggest a role for water system investment within long-term gastric cancer prevention strategies.

## Introduction

Helicobacter pylori (H. pylori) is a common human pathogen with recognized potential for waterborne transmission. Similar to its notorious ability to survive the harsh acidic environment of the gastric epithelium, the organism’s tenacity allows persistence within pipe distribution systems.^1^ In response to stressors such as hyperosmolarity and nutritional deprivation, H. pylori may transition from its spiral morphology to a coccoid viable-but-non-culturable (VBNC) state, allowing survival amidst adverse conditions including chlorine exposure.^2^ H. pylori is also known capable of biofilm formation, another characteristic providing protection from disinfection.^3^

Intermittent water supply (IWS) refers to a non-continuous distribution of water through pipe networks. Such a phenomenon is prevalent in developing regions such as the Caribbean, Latin America, South Asia and African countries, and is estimated to affect more than one billion worldwide.^4^ In resource rich regions, supply intermittence may occur only during times of drought, pollution accident or maintenance, whereas permanent intermittence is often the norm for utility systems in low and middle income countries (LMICs).^5^ Irrespective, IWS, beyond being an inconvenience, poses significant risk of microbiological contamination.^6,7,8^

During periods of supply interruption, pressure within distribution networks drops, facilitating intrusion of surrounding soil and groundwater into pipes through cracks, joints and service connections.^9^ Additionally, water stagnation during non-supply periods may further promote biofilm formation, potentially creating niches favorable for H. pylori persistence.^10^

The contribution of IWS to water pathogen transmission has received limited attention in academic literature however the implications are potentially profound. H. pylori is classified as a WHO/IARC group 1 carcinogen and is responsible for 75-89% of non-cardia gastric cancer.^11^ MALT lymphoma, peptic ulcer disease, iron deficiency anemia and vitamin B12 deficiency represent additional clinically important manifestations contributing substantial global morbidity.^12^

The present study sought to address this gap through a global ecological analysis leveraging utility-level water infrastructure performance data to examine the association between water supply continuity and H. pylori prevalence across countries. Secondary analyses explored whether broader water supply performance indicators including measures of water quality and financial sustainability were associated with H. pylori burden. Identifying modifiable infrastructure determinants of H. pylori transmission may carry important public health implications in view of the substantial global burden associated with gastroduodenal diseases and malignancy.

## Methods

### Data Sources

Water infrastructure data were obtained from the International Benchmarking Network for Water and Sanitation Utilities (IBNET) database, accessed via an expanded utility level export provided by the World Bank IBNET team spanning 1994-2022. IBNET compiles annual performance indicators submitted by water utilities worldwide, with each record representing a single utility-year observation across operational, financial and water quality domains.^13^ Sanitation data were obtained from the WHO/UNICEF Joint Monitoring Programme (JMP) 2025 global database.^14^ Population density data (persons per km2 of land area) were obtained from the World Bank Open Data Indicator.^15^ Country level H. pylori prevalence data were obtained from Chen et al. (Gastroenterology, 2024),^16^ a global meta-analysis including 1748 articles from 111 countries, encompassing 6,270,520 participants spanning 1980-2022.

### Study Design

A cross-sectional ecological study was conducted at the country level. Countries were eligible for inclusion if they appeared in both the IBNET database and the Chen et al. metanalytic prevalence dataset, with at least one utility-year observation for a given indicator. A total of 79 countries had matching data across both sources. The final analytic sample varied by water supply indicator.

### Exposure Variables and Outcome

Four water infrastructure indicators were examined. The primary exposure was continuity of water supply, defined as the mean number of hours per day that water was available at the utility level. Three secondary exposures were also examined: NRW %, calculated as the proportion of produced water not reaching consumers; pipe breaks per utility, defined as the total number of pipe breaks per year reported across the utility’s distribution network; and operating cost coverage ratio, defined as total operating revenue divided by total operating expenses expressed as a percentage. All utility-year observations were aggregated to the country level using unweighted means across all available utility-year records within each country. The outcome was country-level H. pylori prevalence (%), operationalized as the pooled national prevalence estimate derived from Chen et al. using random-effects meta-analysis with multivariable meta-regression adjusting for study period, WHO region, Human Development Index rank and H. pylori detection method.

### Covariates

Two covariates were included in adjusted models. Basic sanitation coverage was defined as the percentage of the national population using at least basic sanitation services, sourced from the JMP 2025 database. This variable was considered a confounder as it captures fecal-oral transmission risk through pathways independent of piped water continuity. JMP sanitation values were averaged across the years corresponding to each country’s available IBNET observations, matching the exposure time window on a country-by-country basis. Log-transformed population density (persons/km2) was included as a second covariate as higher population density may facilitate person-to-person H. pylori transmission independent of water supply infrastructure. Population density data were obtained from the World Bank for the year closest to the midpoint of each country’s IBNET observation window. Candidate covariates including GDP per capita, urbanization rate and age under-5 mortality were evaluated but demonstrated substantial inter-correlation with basic sanitation coverage. Log population density was selected based on conceptual justification, near-zero collinearity with basic sanitation (r = −0.007), and associations with both the exposure and outcome in unadjusted analyses.

### Statistical Analysis

For each exposure variable, two linear regression models were estimated. An unadjusted model regressed H. pylori prevalence on the exposure alone and an adjusted model included basic sanitation coverage and log-transformed population density as covariates. Heteroscedasticity-consistent standard errors were applied in all models. Results are reported as unstandardized regression coefficients (β) with 95% confidence intervals and two-sided p-values. Spearman rank correlations were computed for each exposure-outcome pair to assess the monotonicity of associations independent of linearity assumptions. As a sensitivity analysis, the JMP indicator “water available when needed”; representing the percentage of the national population reporting water availability on demand and averaged across 2021-2024, was examined as an independent, population-based operationalization of IWS. Given the reduced sample available for this indicator (N=55), the sensitivity model was adjusted for basic sanitation coverage only, to preserve adequate degrees of freedom while controlling for the primary confounder of interest. All analyses were conducted in R version 4.5.2.

## Results

### Study Sample

Seventy-nine countries had matching IBNET and H. pylori prevalence data and were included in at least one analysis. The geographic distribution of H. pylori prevalence across all countries with available data, along with the subset of countries contributing IBNET water infrastructure data is displayed in Figure 1. The number of countries included in each regression model varied by indicator availability. 74 countries included continuity of supply and NRW %, 72 contained operating cost coverage and 67 supplied pipe break data. Country-level values for all study variables are provided in Supplementary Table 1.

**Figure 1.**
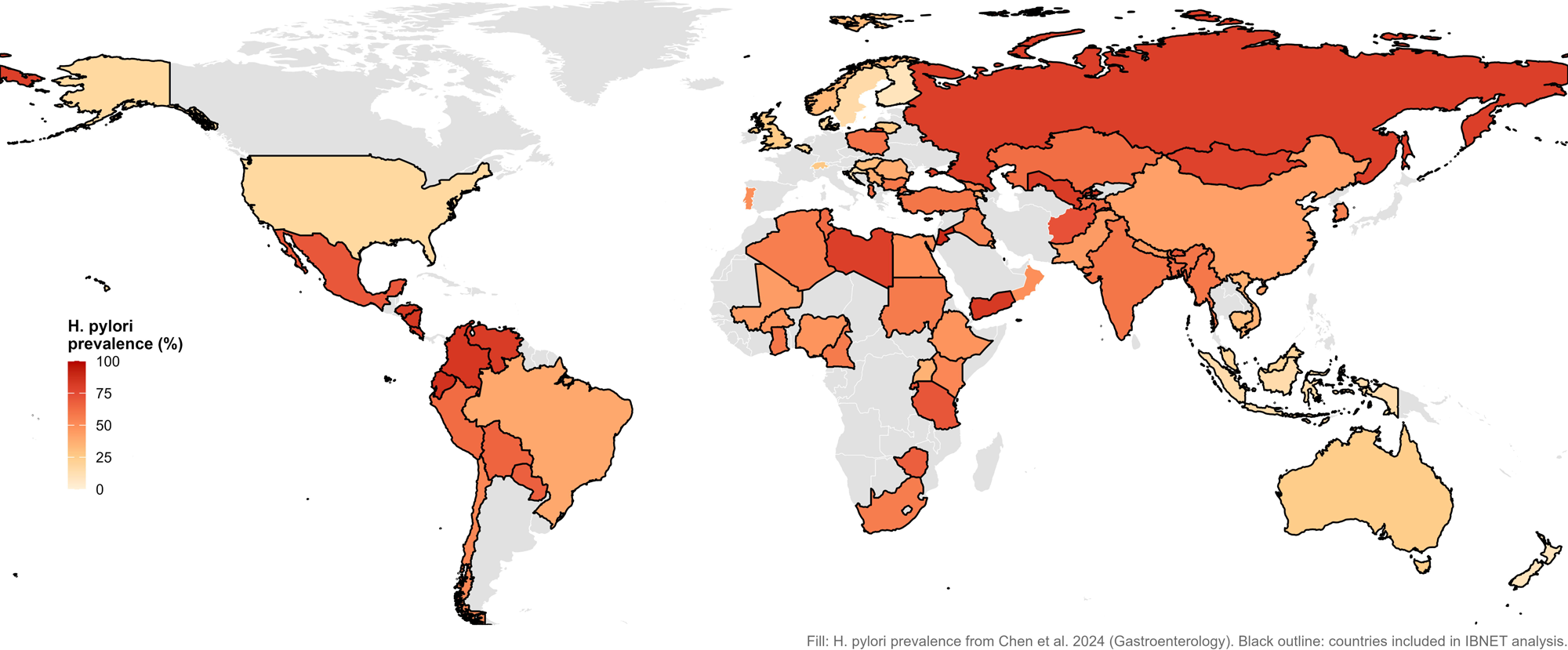
Global distribution of Helicobacter pylori prevalence and countries with IBNET water infrastructure data. Country-level H. pylori prevalence estimates in adults are displayed as a color gradient from low (light) to high (dark red). Countries outlined in black contributed utility-level water infrastructure data from the International Benchmarking Network for Water and Sanitation Utilities (IBNET) database and were included in the primary analysis. Grey countries lacked either H. pylori prevalence data or IBNET data and were excluded. Prevalence estimates were obtained from Chen et al. (Gastroenterology, 2024). IBNET = International Benchmarking Network for Water and Sanitation Utilities.

Mean country-level H. pylori prevalence across the full sample was 51.7% (SD 20.9%), ranging from 9.1% in Finland to 88.6% in Jordan. Prevalence was highest in the Americas (mean 66.4%, SD 19.5%) and Eastern Mediterranean (mean 63.4%, SD 16.5%) regions and lowest in the Western Pacific (mean 37.7%, SD 20.5%) and European (mean 41.5%, SD 20.6%) regions. Mean continuity of supply was 19.7 hours per day (SD 5.7), ranging from 5 hours per day in part of Southeast Asia to 24 hours per day across much of Europe and the Western Pacific. Mean basic sanitation coverage was 73.8% (SD 28%). Descriptive statistics stratified by WHO region are presented in Table 1.

**Table 1.** Country-level descriptive statistics by WHO region. Continuity of supply, non-revenue water percentage, pipe breaks, and operating cost coverage were derived from IBNET utility-level data (World Bank, 1994–2022) and aggregated to the country level using unweighted means. Basic sanitation coverage was obtained from the WHO/UNICEF Joint Monitoring Programme (JMP) 2025 database. Log population density was derived from World Bank population density estimates. H. pylori prevalence estimates were obtained from Chen et al. (Gastroenterology, 2024). IBNET = International Benchmarking Network for Water and Sanitation Utilities.

| Variable | WHO Region | N | Mean (SD) | Median | Range |
| --- | --- | --- | --- | --- | --- |
| H. pylori prevalence (%) | Global | 79 | 51.75 (20.85) | 54.9 | 9.1–88.6 |
|  | African | 13 | 54.79 (10.67) | 56.1 | 36.0–71.1 |
|  | Americas | 14 | 66.39 (19.45) | 69.1 | 17.6–85.7 |
|  | Eastern Mediterranean | 12 | 63.38 (16.49) | 60.3 | 41.3–88.6 |
|  | European | 25 | 41.46 (20.55) | 37.8 | 9.1–81.3 |
|  | Southeast Asia | 6 | 51.60 (19.94) | 58.9 | 14.4–70.1 |
|  | Western Pacific | 9 | 37.73 (20.47) | 36.8 | 9.2–78.8 |
| Continuity of supply (hrs/day) | Global | 74 | 19.72 (5.71) | 23.7 | 4.98–24.0 |
|  | African | 13 | 17.42 (5.00) | 17.1 | 10.1–24.0 |
|  | Americas | 14 | 20.67 (4.78) | 23.1 | 8.48–24.0 |
|  | Eastern Mediterranean | 10 | 17.93 (7.38) | 21.4 | 6.99–24.0 |
|  | European | 22 | 22.14 (4.00) | 24.0 | 10.8–24.0 |
|  | Southeast Asia | 6 | 10.96 (4.91) | 9.89 | 4.98–19.5 |
|  | Western Pacific | 9 | 23.48 (1.05) | 24.0 | 21.3–24.0 |
| Non-revenue water (%) | Global | 74 | 33.50 (14.90) | 30.6 | 0.0–76.6 |
|  | African | 12 | 37.37 (8.94) | 39.4 | 18.3–53.5 |
|  | Americas | 14 | 42.48 (15.60) | 42.5 | 14.2–71.1 |
|  | Eastern Mediterranean | 11 | 33.40 (15.76) | 31.3 | 0.0–54.9 |
|  | European | 23 | 31.69 (15.91) | 25.6 | 6.72–76.6 |
|  | Southeast Asia | 5 | 30.89 (9.15) | 30.6 | 20.3–41.9 |
|  | Western Pacific | 9 | 20.58 (10.35) | 20.9 | 3.93–34.8 |
| Pipe breaks (per utility) | Global | 67 | 3798 (6193) | 926.5 | 0–30536 |
|  | African | 11 | 7156 (10139) | 1739.7 | 793–30536 |
|  | Americas | 12 | 5229 (5235) | 3910.8 | 172–13664 |
|  | Eastern Mediterranean | 10 | 5338 (6809) | 2109.4 | 43–19996 |
|  | European | 22 | 1398 (2181) | 565.1 | 101–10067 |
|  | Southeast Asia | 3 | 1801 (1241) | 2481.5 | 369–2552 |
|  | Western Pacific | 9 | 2608 (6690) | 464.5 | 0–20430 |
| Operating cost coverage (%) | Global | 72 | 130.1 (50.6) | 123.2 | 8.65–271.8 |
|  | African | 12 | 123.0 (37.1) | 119.2 | 53.5–193.5 |
|  | Americas | 14 | 142.7 (40.9) | 151.5 | 82.5–215.2 |
|  | Eastern Mediterranean | 10 | 78.1 (45.0) | 80.4 | 8.65–179.6 |
|  | European | 23 | 136.3 (50.3) | 125.2 | 58.5–271.8 |
|  | Southeast Asia | 5 | 113.8 (30.6) | 122.9 | 60.6–136.0 |
|  | Western Pacific | 8 | 176.2 (50.7) | 177.9 | 107.4–265.2 |
| Basic sanitation coverage (%) | Global | 77 | 74.12 (27.99) | 84.9 | 6.34–100.0 |
|  | African | 13 | 35.41 (23.18) | 33.3 | 6.34–84.5 |
|  | Americas | 13 | 80.53 (15.21) | 81.5 | 39.6–99.7 |
|  | Eastern Mediterranean | 12 | 75.89 (28.63) | 90.6 | 22.9–100.0 |
|  | European | 25 | 94.11 (6.62) | 97.3 | 77.6–99.9 |
|  | Southeast Asia | 5 | 45.20 (7.11) | 46.5 | 34.0–53.4 |
|  | Western Pacific | 9 | 78.92 (26.54) | 94.0 | 25.8–100.0 |
| Log population density | Global | 79 | 4.26 (1.35) | 4.37 | 0.68–8.95 |
|  | African | 13 | 4.31 (0.97) | 4.26 | 2.76–6.18 |
|  | Americas | 14 | 3.73 (0.79) | 3.67 | 2.33–5.39 |
|  | Eastern Mediterranean | 12 | 4.31 (1.56) | 4.39 | 1.33–7.48 |
|  | European | 25 | 4.25 (1.00) | 4.45 | 1.92–5.92 |
|  | Southeast Asia | 6 | 5.13 (1.43) | 5.11 | 2.97–7.12 |
|  | Western Pacific | 9 | 4.41 (2.57) | 4.57 | 0.68–8.95 |

IBNET = International Benchmarking Network for Water and Sanitation Utilities. SD = standard deviation.

### Primary Analysis: Continuity of Water Supply

Greater continuity of water supply was associated with lower H. pylori prevalence in both unadjusted and adjusted models (Table 2, Figure 2). Each additional hour per day of water supply was associated with a 0.99% lower H. pylori prevalence in the unadjusted model (β = −0.987, 95% CI −1.669 to −0.305, p = 0.005, N = 74). After adjustment for basic sanitation coverage and log population density, the association strengthened and remained statistically significant (β = −1.125, 95% CI −1.876 to −0.375, p = 0.004, N = 72). Figure 3 displays the partial regression plot, illustrating that the inverse association between water supply continuity and H. pylori prevalence was preserved after removing the shared influence of sanitation coverage and population density from both variables. Spearman rank correlation confirmed the expected negative direction of association (ρ = −0.321, p = 0.005; Supplementary Table 2).

**Figure 2.**
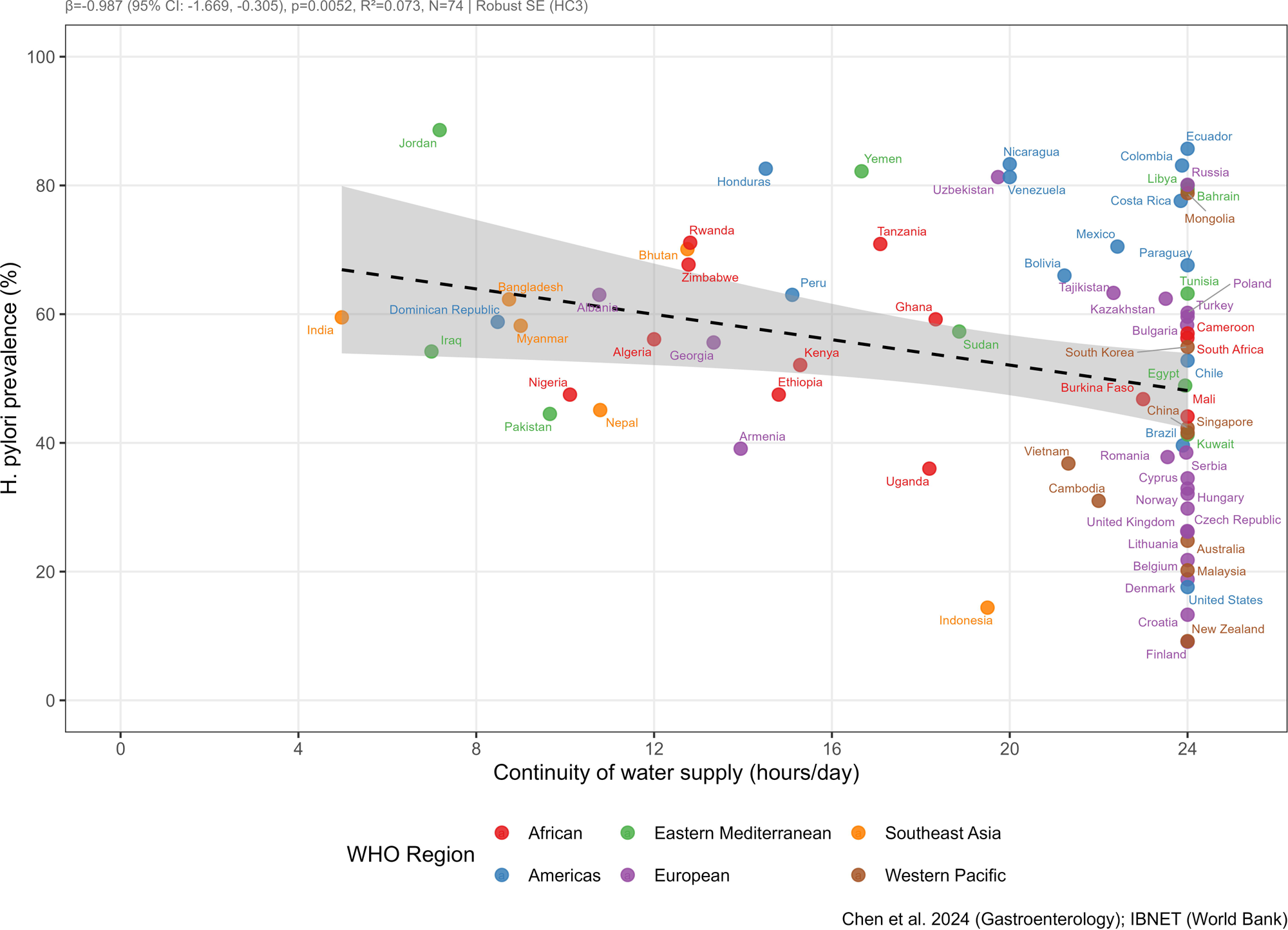
Association between continuity of water supply and Helicobacter pylori prevalence. Scatterplot displaying the unadjusted relationship between mean continuity of water supply (hours per day) and country-level H. pylori prevalence (%). Each point represents one country (N = 74). The fitted regression line with 95% confidence interval is shown. Greater water supply continuity was associated with significantly lower H. pylori prevalence (β = −0.987, 95% CI −1.669 to −0.305, p = 0.005).

**Figure 3.**
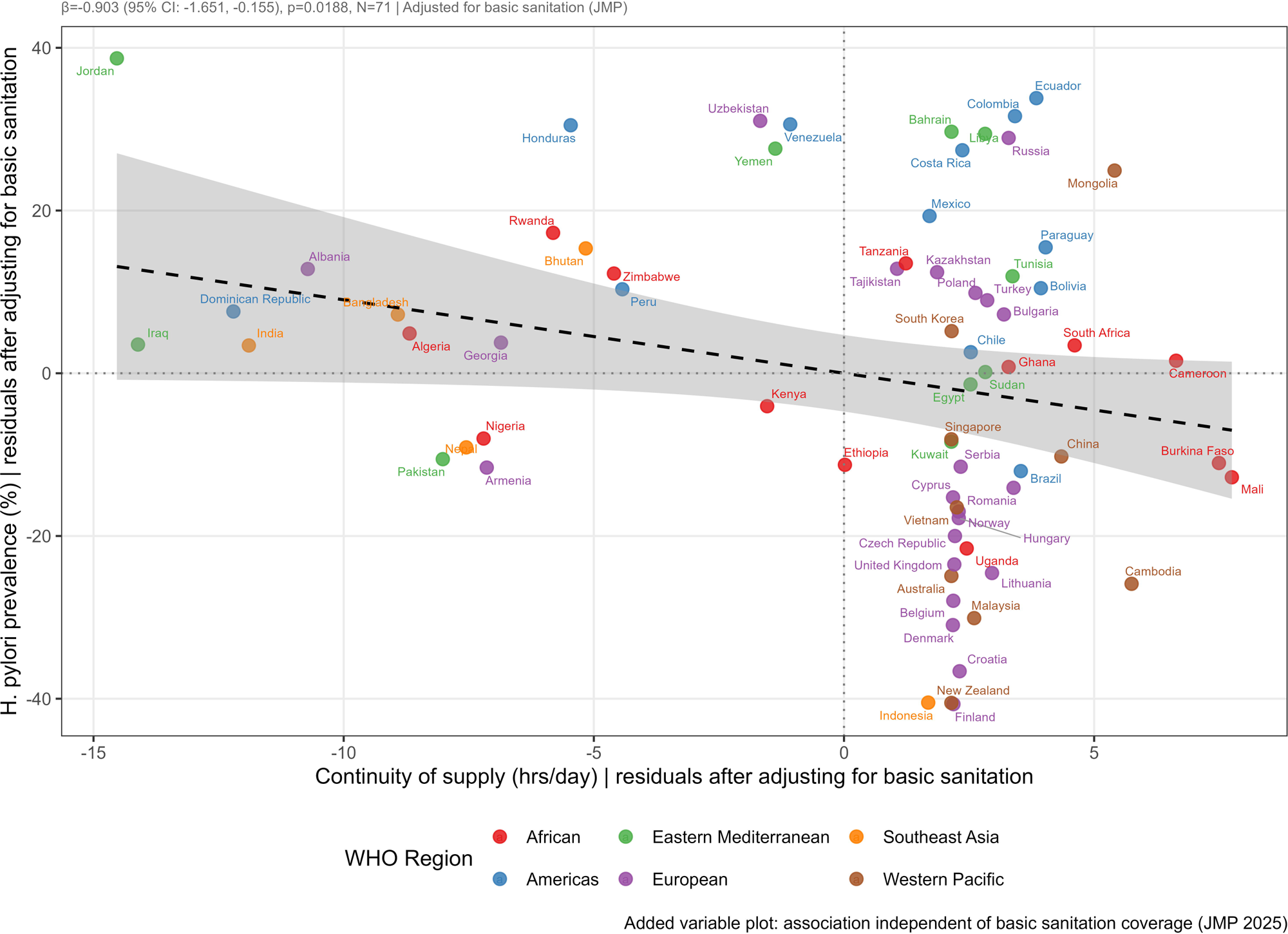
Partial regression plot of the association between continuity of water supply and Helicobacter pylori prevalence, adjusted for basic sanitation coverage and log population density. The plot displays the independent association between continuity of water supply and H. pylori prevalence after removing the shared influence of basic sanitation coverage and log-transformed population density from both variables. Each point represents one country (N = 72). The fitted regression line with 95% confidence interval is shown. The inverse association between water supply continuity and H. pylori prevalence was preserved after adjustment (β = −1.125, 95% CI −1.876 to −0.375, p = 0.004).

**Table 2.** Association between water infrastructure indicators and H. pylori prevalence: unadjusted and sanitation– and population density-adjusted regression models. Panel A presents unadjusted linear regression models. Panel B presents models adjusted for basic sanitation coverage (WHO/UNICEF JMP) and log-transformed population density (World Bank). The outcome in all models was country-level *H. pylori* prevalence (%) derived from Chen et al. (*Gastroenterology*, 2024). Heteroskedasticity-consistent standard errors (HC3) were applied throughout. β = unstandardized regression coefficient; CI = confidence interval; R² = coefficient of determination.

| Tier | Variable | N | $\beta$ | 95% CI | p-value | R<sup>2</sup> |
| --- | --- | --- | --- | --- | --- | --- |
| Primary | Continuity of supply (hrs/day) | 74 | -0.987 | -1.669 to -0.305 | 0.005 | 0.073 |
| Secondary | Non-revenue water (%) | 74 | 0.644 | 0.301 to 0.987 | <0.001 | 0.216 |
| Secondary | Pipe breaks (per utility) | 67 | 0.001 | 0.000 to 0.002 | 0.161 | 0.039 |
| Secondary | Operating cost coverage (%) | 72 | -0.115 | -0.206 to -0.024 | 0.014 | 0.077 |

**Panel B: Adjusted for basic sanitation coverage and log population density**
| Tier | Variable | N | $\beta$ | 95% CI | p-value | R<sup>2</sup> |
| --- | --- | --- | --- | --- | --- | --- |
| Primary | Continuity of supply (hrs/day) | 72 | -1.125 | -1.876 to -0.375 | 0.004 | 0.111 |
| Secondary | Non-revenue water (%) | 73 | 0.620 | 0.282 to 0.958 | <0.001 | 0.243 |
| Secondary | Pipe breaks (per utility) | 66 | 0.000 | -0.001 to 0.002 | 0.354 | 0.049 |
| Secondary | Operating cost coverage (%) | 71 | -0.109 | -0.196 to -0.022 | 0.015 | 0.105 |

$\beta$ = unstandardized regression coefficient. HC3 heteroskedasticity-consistent standard errors. R<sup>2</sup> = coefficient of determination.

### Secondary Analyses

Results for all secondary exposure variables are presented in Table 2 and displayed in Figure 4. Spearman correlations for all exposure variables are presented in Supplementary Table 2. NRW % was positively associated with H. pylori prevalence in both unadjusted and adjusted models. Each percentage point increase in non-revenue water was associated with a 0.64% higher H. pylori prevalence in the unadjusted model (β = 0.644, 95% CI 0.301 to 0.987, p < 0.001, N = 74). This association was robust to adjustment for basic sanitation coverage and log population density (β = 0.620, 95% CI 0.282 to 0.958, p < 0.001, N = 73). Spearman correlation was in the expected positive direction (ρ = 0.484, p < 0.001).

**Figure 4.**
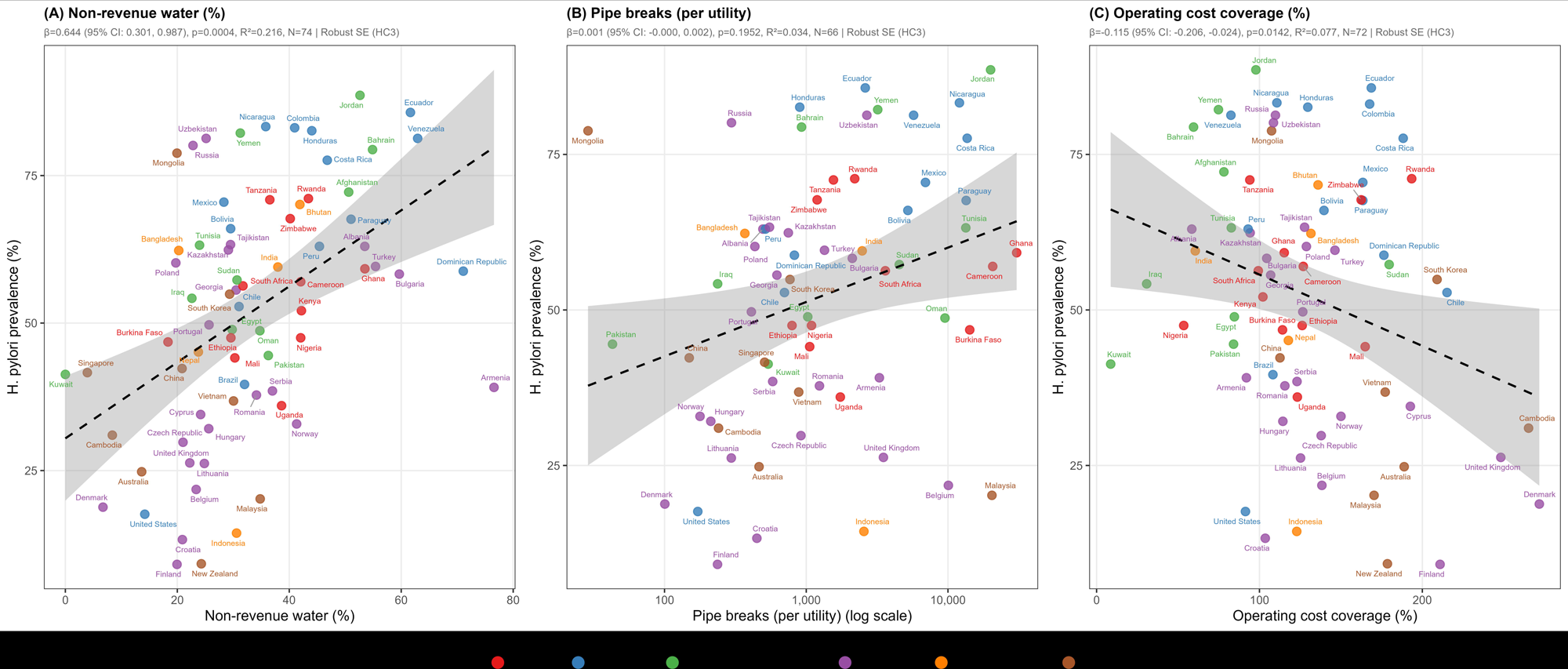
Unadjusted associations between secondary water infrastructure indicators and Helicobacter pylori prevalence. Scatterplots displaying unadjusted associations between country-level H. pylori prevalence (%) and three secondary water infrastructure indicators: non-revenue water percentage (NRW%; N = 74), pipe breaks per utility (N = 67), and operating cost coverage ratio (N = 72). Each point represents one country. Fitted regression lines with 95% confidence intervals are shown. Higher NRW% was positively associated with H. pylori prevalence (β = 0.644, p < 0.001); higher operating cost coverage was inversely associated (β = −0.115, p = 0.014); pipe breaks per utility were not significantly associated in regression models (β = 0.001, p = 0.195). NRW = non-revenue water.

Operating cost coverage was inversely associated with H. pylori prevalence in both the unadjusted (β = −0.115, 95% CI −0.206 to −0.024, p = 0.014, R² = 0.077, N = 72) and adjusted models (β = −0.109, 95% CI −0.196 to −0.022, p = 0.015, N = 71). The magnitude of the association was largely unchanged after adjustment. Spearman correlation was in the expected negative direction (ρ = −0.236, p = 0.046).

Pipe breaks per utility were not significantly associated with H. pylori prevalence in either the unadjusted (β = 0.001, 95% CI 0 to 0.002, p = 0.195, N = 67) or adjusted model (β = 0.000, 95% CI −0.001 to 0.002, p = 0.354, N = 66). The pipe breaks distribution was markedly right-skewed, with a mean of 3,798 breaks per utility (SD 6,193) and values ranging from 0 to 30,536, reflecting substantial heterogeneity in utility size and reporting practices across countries. Despite the null regression result, the Spearman correlation was in the expected positive direction (ρ = 0.368, p = 0.002).

### Sensitivity Analysis

When intermittent water supply was operationalized using the JMP population-based measure of water availability on demand, findings were consistent with the primary IBNET-based analysis (Supplementary Figure 1). A higher percentage of the population reporting water available when needed was associated with lower H. pylori prevalence in the unadjusted model (β = −0.277, 95% CI −0.494 to −0.060, p = 0.013, R² = 0.079) and the association strengthened after adjustment for basic sanitation coverage (β = −0.434, 95% CI −0.673 to −0.194, p < 0.001, R² = 0.145). The Spearman correlation was in the expected negative direction (ρ = −0.266, p = 0.050).

## Discussion

Despite substantial epidemiological evidence implicating drinking water distribution supplies in H. pylori transmission, the contribution of supply intermittency to population-level infection burden has not previously been examined on a global scale. This study addresses this gap through a cross-sectional ecological analysis of 79 countries. The primary finding indicate greater water supply discontinuity to be associated with increased H. pylori prevalence, with persistent association after adjustment for basic sanitation coverage and log population density. Higher NRW % and lower operating cost coverage were each independently associated with higher H. pylori prevalence, consistent with the hypothesis that broader infrastructure deterioration contributes to pathogen transmission risk. Pipe breaks per utility were not significantly associated with H. pylori prevalence in regression models, though the Spearman correlation was in the expected direction.

### Intermittent Water Supply and H. pylori

The inverse association between water supply continuity and H. pylori prevalence observed builds upon a prior body of literature implicating piped water distribution as a vehicle for H. pylori transmission. Amongst the earliest evidence, Klein et al. demonstrated that Peruvian children supplied with municipal water were significantly more likely to be infected with H. pylori than those sourcing from community wells. This association persisted after socioeconomic adjustment suggesting the distribution system itself as contributory to infection risk.^17^ H. pylori has been identified in 15.7% of drinking water sample globally, with detection reported across all world regions and income levels.^18^ The present findings expand the evidence for waterborne transmission of H pylori to the infrastructure level, illustrating hours of daily water supply as a meaningful correlate of H. pylori prevalence at the country level.

The mechanistic pathways linking water distribution infrastructure to H. pylori transmission have been conceptually modeled through several interconnected processes. Pressure-driven intrusion during supply interruptions, stagnation-associated biofilm accumulation and persistence of the organism in a VBNC state are all likely contributory.^19^

Experimental studies demonstrate that IWS conditions directly alter the microbial ecology of distribution-system biofilms. Following restoration of flow, accumulated biofilm material and associated microorganisms are mobilized in bulk to consumers. In a controlled pipe-facility model, Preciado and colleagues showed that prolonged stagnation during supply interruptions altered the composition of pipe biofilms, favoring the growth of pathogenic organisms over commensal species. This effect worsened with longer interruptions.^20^

H. pylori further demonstrates features facilitating environmental persistence. The VBNC state is associated with resistance to standard chlorination practices.^21^ Within biofilms, resistance is amplified as biofilm embedded organisms exhibit substantially greater disinfectant tolerance than planktonic forms.^22^

Together these mechanisms provide a coherent biological basis for the observed ecological association between IWS and H. pylori prevalence.

### Secondary Findings

The secondary findings are best understood as complementary markers of overall water system deterioration. NRW % reflects the proportion of produced water lost before reaching consumers and was positively associated with H. pylori prevalence, with an unadjusted R² of 0.216, robust to adjustment for sanitation coverage and population density. High NRW % is indicative of aging or deteriorating pipe networks and represents the same structural conditions permitting microbial ingress. Operating cost coverage, a measure of financial sustainability, was also inversely associated with H. pylori prevalence and its coefficient was largely unchanged after sanitation adjustment suggesting that utilities operating at a financial deficit may be systematically less able to maintain supply continuity or sustain disinfection practices. Interpreted together, these findings suggest that the relationship between water infrastructure and H. pylori prevalence is unlikely to represent a single deficiency but rather, a cumulative microbiological consequence of chronic infrastructure deterioration and underinvestment in water distribution systems.

Pipe breaks per utility did not reach statistical significance in either regression model, though the Spearman rank correlation was in the expected positive direction (ρ = 0.368, p = 0.002). This discordance may reflect the measurement heterogeneity inherent to this variable rather than a true absence of association. The raw pipe break distribution was markedly right-skewed (mean 3,798, SD 6,193, range 0 to 30,536 per utility), driven by variation in utility size, geographic coverage, and reporting completeness across countries. Raw break counts are not normalized to network length or connection density in the available IBNET data, making meaningful cross-country comparison difficult. The Spearman correlation, being rank-based and therefore less sensitive to extreme values, is likely an accurate reflection of the relationship. The persistence of association between IWS and H. pylori prevalence within the sensitivity analysis using an independent population-based measure of water availability suggests a generalizable pattern rather than a feature of any single data source.

### Limitations

Several limitations were identified. The ecological design precludes causal inference. Since country-level estimates were used for both exposure and outcome, findings reflect national-level associations and cannot be extrapolated to subnational regions, individual utilities or persons.

Country-level aggregation of IBNET utility data obscures potentially important within-country heterogeneity in infrastructure quality, and countries with more developed utility reporting systems may be systematically overrepresented in the dataset, introducing selection bias. IBNET indicators are self-reported by utilities and subject to variable data quality. H. pylori prevalence estimates derived from the Chen et al. meta-analysis reflect variable study designs, diagnostic methods, and time periods across contributing countries, and country-level estimates carry uncertainty not fully captured by point estimates alone. Residual confounding by factors not available in the current dataset cannot be excluded. The cross-sectional design precludes assessment of temporality, and temporal alignment between utility performance data and prevalence estimates could not be assured for all countries.

## Conclusion

This global ecological study identified a consistent direct association between IWS and H. pylori prevalence across countries and demonstrated replicability using an independent population-based measure of water availability. Secondary associations involving NRW % and operating cost coverage further suggest that broader deterioration of water infrastructure and utility sustainability may contribute meaningfully to transmission risk. Collectively, these findings support the hypothesis that IWS is not merely a marker of underdevelopment, but a potentially relevant environmental determinant within the epidemiology of H. pylori infection. Beyond the immediate implications for H. pylori transmission, this analysis frames water distribution networks as biologically active systems capable of shaping population exposure to H. pylori. In regions where H. pylori prevalence and gastric cancer burden remain high, durable reduction in disease incidence may require approaches extending beyond test-and-treat strategies. Investments in reliable, continuous, and microbiologically secure water infrastructures represent an underrecognized component of long-term gastric cancer prevention strategy and global gastrointestinal health.

## Data Availability

The H. pylori prevalence data used in this study are derived from Chen et al. (Gastroenterology, 2024) and are available from the corresponding author upon reasonable request. Water infrastructure data were obtained from IBNET, the World Bank (https://newibnet.org) and sanitation data from the WHO/UNICEF Joint Monitoring Programme (https://washdata.org), both publicly accessible. Population density data are available via the World Bank Open Data platform (https://data.worldbank.org).

https://newibnet.org

## Acknowledgements

The authors received no financial support for the research, authorship, or publication of this article. The authors thank Alexander Danilenko (IBNET, the World Bank) for providing the expanded utility-level data export used in this analysis.

## Ethics Statement

This study used only publicly available, aggregated country-level data from the World Bank IBNET database, the WHO/UNICEF Joint Monitoring Programme, and a published meta-analysis. No individual patient data, biological specimens, or identifiable information were used. Accordingly, this study was exempt from institutional review board oversight.

## Author Contributions

SSG: conceptualization, data curation, formal analysis, writing – original draft. CW, BT, MJW: writing – review and editing. DT: methodology, supervision, writing – review and editing.

## References

1. Park SR, Mackay WG, Reid DC. Helicobacter sp. recovered from drinking water biofilm sampled from a water distribution system. Water Res. 2001;35(6):1624–1626. doi:10.1016/s0043-1354(00)00582-0

2. Moreno Y, Piqueres P, Alonso JL, Jiménez A, González A, Ferrús MA. Survival and viability of Helicobacter pylori after inoculation into chlorinated drinking water. Water Res. 2007;41(15):3490–3496.

3. Moreno Y, Moreno-Mesonero L, Soler P, Zornoza A, Soriano A. Influence of drinking water biofilm microbiome on water quality: Insights from a real-scale distribution system. Sci Total Environ. 2024;921:171086. doi:10.1016/j.scitotenv.2024.171086

4. Kumpel E, Billava N, Nayak N, Ercumen A. Water use behaviors and water access in intermittent and continuous water supply areas during the COVID-19 pandemic. J Water Health. 2022;20(1):139–148. doi:10.2166/wh.2021.184

5. Galaitsi SE, Russell R, Bishara A, Durant JL, Bogle J, Huber-Lee A. Intermittent domestic water supply: A critical review and analysis of causal-consequential pathways. Water. 2016;8(7):274.

6. Kumpel E, Nelson KL. Comparing microbial water quality in an intermittent and continuous piped water supply. Water Res. 2013;47(14):5176–5188. doi:10.1016/j.watres.2013.05.058

7. Bivins A, Lowry S, Wankhede S, et al. Microbial water quality improvement associated with transitioning from intermittent to continuous water supply in Nagpur, India. Water Res. 2021;201:117301. doi:10.1016/j.watres.2021.117301

8. van den Berg H, Quaye MN, Nguluve E, Schijven J, Ferrero G. Effect of operational strategies on microbial water quality in small scale intermittent water supply systems: The case of Moamba, Mozambique. Int J Hyg Environ Health. 2021;236:113794. doi:10.1016/j.ijheh.2021.113794

9. Taylor DDJ, Slocum AH, Whittle AJ. Analytical scaling relations to evaluate leakage and intrusion in intermittent water supply systems. PLoS One. 2018;13(5):e0196887. doi:10.1371/journal.pone.0196887

10. Gião MS, Azevedo NF, Wilks SA, Vieira MJ, Keevil CW. Persistence of Helicobacter pylori in heterotrophic drinking-water biofilms. Appl Environ Microbiol. 2008;74(19):5898–5904. doi:10.1128/AEM.00827-08

11. Plummer M, Franceschi S, Vignat J, Forman D, de Martel C. Global burden of gastric cancer attributable to Helicobacter pylori. Int J Cancer. 2015;136(2):487–490.

12. Robinson K, Atherton JC. The Spectrum of Helicobacter-Mediated Diseases. Annu Rev Pathol. 2021;16:123–144. doi:10.1146/annurev-pathol-032520-024949

13. World Bank. International Benchmarking Network for Water and Sanitation Utilities (IBNET). https://newibnet.org/about-us-0. Accessed May 7, 2026.

14. World Health Organization/UNICEF Joint Monitoring Programme for Water Supply, Sanitation and Hygiene (JMP). Estimates for Drinking Water, Sanitation and Hygiene Services by Country (2000–2024). 2025. https://washdata.org/data/country/WLD/household/download. Accessed May 7, 2026.

15. World Bank. Population density (people per sq. km of land area) [EN.POP.DNST]. World Bank Open Data. https://data.worldbank.org/indicator/EN.POP.DNST. Accessed May 2026.

16. Chen YC, Malfertheiner P, Yu HT, et al. Global prevalence of Helicobacter pylori infection and incidence of gastric cancer between 1980 and 2022. Gastroenterology. 2024;166(4):605–619.

17. Klein PD, Opekun AR, Smith EO, Graham DY, Gaillour A; Gastrointestinal Physiology Working Group. Water source as risk factor for Helicobacter pylori infection in Peruvian children. Lancet. 1991;337(8756):1503–1506.

18. Ekundayo TC, Swalaha FM, Ijabadeniyi OA. Global and regional prevalence of Helicobacter pylori in drinking waters: A sustainable, human development and socio-demographic indices based meta-regression-modelling. Sci Total Environ. 2023;861:160633. doi:10.1016/j.scitotenv.2022.160633

19. Bellack NR, Koehoorn MW, MacNab YC, Morshed MG. A conceptual model of water’s role as a reservoir in Helicobacter pylori transmission: a review of the evidence. Epidemiol Infect. 2006;134(3):439–449.

20. Calero Preciado C, Husband S, Boxall J, Del Olmo G, Soria-Carrasco V, Maeng SK, Douterelo I. Intermittent water supply impacts on distribution system biofilms and water quality. Water Res. 2021;201:117372.

21. García A, Salas-Jara MJ, Herrera C, González C. Biofilm and Helicobacter pylori: from environment to human host. World J Gastroenterol. 2014;20(19):5632.

22. Morgan DR, Torres J, Sexton R, et al. Risk of recurrent Helicobacter pylori infection 1 year after initial eradication therapy in 7 Latin American communities. JAMA. 2013;309(6):578–586.

