## Supplementary figures and images for "Association Between Intermittent Water Supply and Helicobacter pylori Prevalence: A Global Ecological Study"

### Supplemental Figure 1

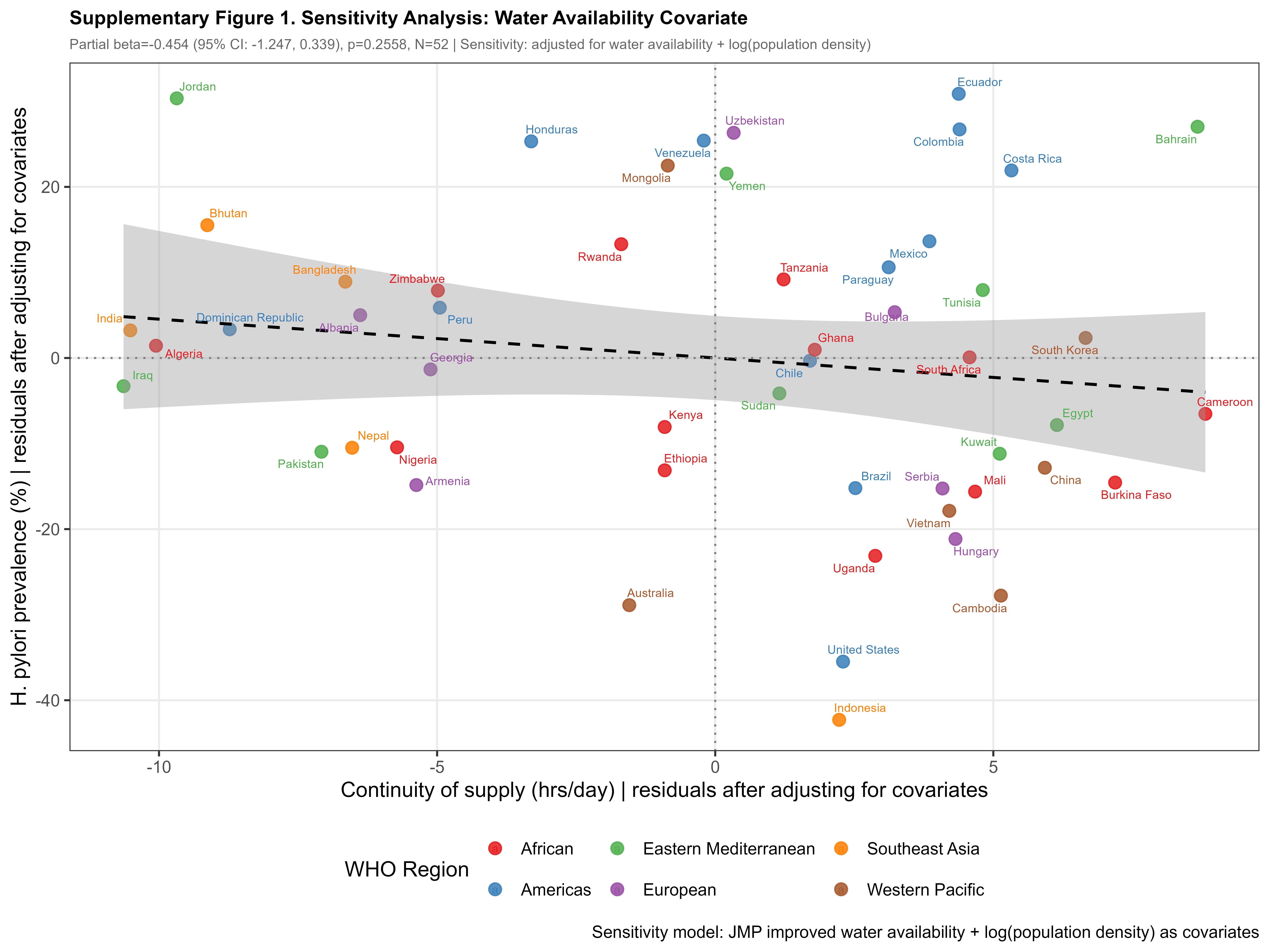
